# Anxiety among the general population during Coronavirus-19 Disease in Saudi Arabia: Implications for a Mental Support Program

**DOI:** 10.1101/2020.05.07.20090225

**Authors:** Salman H. Alsaqri, Mohannad J. Alkwiese, Mohammed K. Aldalaykeh, Mada Ibrahim Hamzi, Mada Mousa Mahdi, Zainab M. Shafie

## Abstract

**Background:** The 2019 outbreak of coronavirus disease (COVID-19) is an worldwide health emergency that threatens mental health of general public. Research data are required to establish evidence-based approaches to alleviate the symptoms of anxiety during the outbreak. This study aimed to survey the general public in Saudi Arabia to help in understanding their levels of anxiety during the COVID-19 outbreak.

**Methods:** The researchers performed an on-line survey using snowball sampling methods from April 1 through April 10, 2020. The online survey collected demographics information data, while the Social Anxiety Questionnaire for Adults (SAQ-A30) was used to measure anxiety level.

**Results:** This study consisted of 709 respondents from different regions of Saudi Arabia. The general population reported mild to moderate rates of anxiety. Married respondents were associated with a significantly higher level of anxiety (p<0.05).

**Conclusions:** The respondents reported mild to moderate anxiety during the outbreak of COVID-19 in Saudi Arabia.

**Implication for practice:** The results of this study identify the implications to lower levels of anxiety that may be used during the COVID-19 outbreak to develop mental health support programs to improve the mental health of vulnerable groups.

## Introduction

The Chinese city of Wuhan has documented a novel pneumonia caused by coronavirus disease 2019 (COVID-19) from end of December 2019 (COVID-19), which is spreading nationally and internationally (Li et al., 2020). The virus has been known as severe acute respiratory syndrome coronavirus 2 (SARS-CoV-2). According to data published by the National Health Commission of China (NHC), the number of reported cases in Chinese mainland rose to 80, 151 as of 2 March 2020 (Zhao et al., 2020) and confirmed cases have been identified in more than a hundred countries. In addition, transmission from one person to another outside China was reported (Rothe et al., 2020). According to Saudi Center for Disease Prevention and Control (SCDC) the number of confirmed cases rose to 27.011 on 4 May 2020 (SCDC, 2020). The World Health Organization (WHO) held an emergency meeting on 30 January 2020 and declared a worldwide outbreak of COVID-19 and a global public health emergency (WHO, 2020).

The continuing COVID-19 outbreak is triggering anxiety and a prompt awareness of the mental health status of society is urgently needed (Xiang et al., 2020). Earlier research has recognized a significant and broad range of psychosocial effects on individuals, populations and globally during infection outbreaks. Individually, people are likely to be fearful of becoming ill or dying, feelings of helplessness and stigma (Hall, Hall, & Chapman, 2008). During one outbreak of influenza, about 10 to 30 % of the general population is highly or extremely anxious about the risk of getting the virus (Rubin, Potts, & Michie, 2010). With schools and businesses closed, the negative emotions felt by people are intensified (Van Bortel et al., 2016). During the SARS epidemic, many studies examined the non-infected population’s mental wellbeing, exposing major mental symptoms such as anxiety that were found to be correlated with younger age and growing self-blame (Sim, Chan, Chong, Chua, & Soon, 2010).

There is currently no known data on the level of public anxiety at the peak of the outbreak of COVID-19. Most of the epidemic-related studies focus on the identification of epidemiology and clinical features of infected people (Chen et al., 2020), the genomic description of the virus (Lu et al., 2020), and global health policy challenges (Rubin & Wessely, 2020). However, there are no studies exploring the mental effects of COVID-19 on Saudi Arabia’s general population. This study thus constitutes the first mental health survey carried out in the general population of Saudi Arabia in the light of the COVID-19 outbreak. The aim of this study is to determine the levels of anxiety that may assist government agencies and healthcare professionals in protecting the public’s mental well-being in the face of a widespread COVID-19 in Saudi Arabia and other regions of the world.

## Methods

### Sample and setting

This study was based on a descriptive, cross-sectional design for assessing the level of public anxiety during the COVID-19 outbreak using an anonymous online survey. A snowball sampling technique was used, which is based on recruiting the general public living in Saudi Arabia during the COVID-19 outbreak. The online survey was initially distributed to students at universities, and promoted others in the general public to compare the inter-regional differences in anxiety among the population in Saudi Arabia, with snowball samples reflecting diverse geographical settings in Saudi Arabia participating in the study. The collection of data lasted 11 days from April 1 to April 10, 2020. The total recorded cases of COVID-19 in Saudi Arabia during this time surpassed 2000 cases. In order to conduct a power analysis for this study, the G^*^power software was used to suppose sample size, suppose 0.95 power with an alpha error of 0.05 and a medium effect size of (0.5), the total sample size will require 700 subjects to have confidence in the findings.

### Data collection process

Since the Saudi Government suggested that the population evade face-to-face interaction and separate themselves at home, prospective respondents were invited by current study respondents electronically. They accomplished the survey through an online survey platform (Google Forms). The survey broken down into sections. The first section includes the cover letter form and the consent form and the demographic sheet describing the purpose and procedures of the study, while the second section includes the social Anxiety Questionnaire (SAQ-A30). Acceptance of expedited ethics was received from the Institutional Review Board (IRB) of the University of Hail (H-2020-080). Announcement on this study has been posted on the devoted university website.

### Survey Development

The structured survey was composed of questions covering a variety of areas: (1) demographic data form that involved questions on the disclosure of age, gender, and marital status that were examined in order to assess their relationship to the anxiety level. (2) Social Anxiety Questionnaire for Adults (SAQ-A30) was established by Caballo et al. (2010). The questionnaire is intended to measure specific and/or generalized social phobia/anxiety in adults population (≥18 years) from general and clinical populations. This is very valuable for recognizing not only people with generalized social phobia but mainly specific social phobia (Caballo et al., 2010). The SAQ-A30 comprises 30 items conforming a social phobia/anxiety structure of five very solid dimensions, each of them including six items. Data on the questionnaire’s internal consistency, construct validity, cut-off scores, invariance, and factor structure have been presented (Caballo et al., 2010). Internal consistency reliability was good for SAQ-A30 in this study (Cronbach’s α = 0.91).

### Ethical considerations

Standing Committee for Research Ethics (SCRE) - University of Hail has reviewed and granted ethical permission for the study. Prior to the start of data collection, the objectives, benefits and risks of the study were addressed with the participants and it was ensured that participation in the study was voluntary. In addition, all participants were made aware that the information would be presented in a group format and that anonymity and confidentiality were assured. Participants were allowed to terminate the survey at any time they desired. Participants will be informed that they may not directly benefit from their participation in the study, but that the findings of the study may be used to develop mental assistance programs can help in alleviating level of anxiety. The risks associated with participating in this study are considered to be minimal. Inconvenience in studying subjects will include time and energy spent during data collection.

### Statistical Analysis

The Statistical Package for Social Sciences (SPSS) software (IBM SPSS Statistics 25) was used to describe the study sample characteristics and determine the mean level of SAQ-A30. For all statistical tests, levels of statistically significant were established at α < 0.05. Independent t-test was used to examine the difference in the mean score of SAQ-A30 between male and female, and between married and unmarried. One-way variance analysis (ANOVA) followed by Scheffe’s post-hoc test was used to examine differences in in mean score of SAQ-A30 between different age groups. The Pearson’s correlation coefficient was used to examine the relationship between the anxiety level of respondents and their age in years.

## Results

### Demographic characteristics of the Respondents

Researchers received 812 answers and 103 respondents did not complete the survey. Researchers eventually included 709 respondents from different regions of Saudi Arabia who completed the survey (completion rate: 92.79%). Respondents were mainly young-aged (mean = 33.29 years, SD 8.68) and their age ranged between 20–68 years old. More than half of them were male (59.7 %), while most of the respondents were married (69.3%). Most of them had primary and secondary education (58.2%), and more than half of them have been employed (56.4%) (Table 1).

Results analysis showed the mean (SD) level of anxiety among Saudi population was 67.2 (2.24) (range = 30-138), measured using the SAQ-A30. The possible score range for this scale is between 30 and 150, with a higher score reflecting a higher level of anxiety.

**Table 1.** Demographic characteristics of the respondents (n = 709)

| Variable | Frequency | % |
| --- | --- | --- |
| **Gender** |  |  |
| Male | 423 | 59.7 |
| Female | 286 | 40.3 |
| **Marital status** |  |  |
| Married | 491 | 69.3 |
| Unmarried | 218 | 30.7 |
| **Age** |  |  |
| Young | 428 | 60.4 |
| Middle age | 269 | 37.9 |
| Elder | 12 | 1.7 |

### Relationship between Demographic Characteristics and Anxiety Level

There was a statistically significant difference between the anxiety level and the marital status (t= −3.63, p =.000) in which married (mean= 69.02) respondents had higher anxiety level than unmarried group (Table 2). However, the results showed no correlation between anxiety level and age in years. Additionally, there were no statistically significant differences between men and women in their perception of anxiety (t= −.601, p=0.54) (Table 2).

**Table 2.** Difference Between Selected Demographics and Anxiety Level (n = 709)

| Variables | n | Mean | SD | F/t | df1 | df2 | p |
| --- | --- | --- | --- | --- | --- | --- | --- |
| **Age** |  |  |  |  | 2 | 706 | .14 |
| Young | 428 | 66.5 | 20.0 | 1.92 |  |  |  |
| Middle age | 269 | 68.7 | 18.4 |  |  |  |  |
| Old | 12 | 60.0 | 15.1 |  |  |  |  |
| **Marital status** |  |  |  |  |  |  |  |
| Married | 491 | 69.0 | 18.6 | -3.6 | 707 |  | .000* |
| Unmarried | 218 | 63.3 | 20.5 |  |  |  |  |
| **Gender** |  |  |  |  |  |  |  |
| Male | 423 | 66.9 | 19.46 | - 0.60 | 707 |  | .54 |
| Female | 286 | 67.8 | 19.39 |  |  |  |  |

## Discussion

The results of this study provide us with an overview of the level of anxiety among the general Saudi population due to the COVID-19 outbreak in the country, and after the WHO declared a state of emergency of international concern for people’s health. The results of this study indicated that the general public in Saudi Arabia reported anxiety levels ranging from mild to moderate. One potential explanation is that Saudi Arabia, as one of the countries of the Middle East, has its social systems, cultural norms and relationships that can vary from the norms of Western cultures. Families and friends in Saudi culture can be instrumental in alleviating tension and anxiety among the general public experiencing COVID-19 outbreaks. According to Pew research center (PRC), the most of Saudi Arabia’s people (93%) were Muslims (PRC, 2018), the sample of this study may depend on the beliefs of individuals in God (Allah), that everything that occurs to a Muslim is by Allah and He alone saves us. God says in Quran: ‘‘It is for those who believe, a guide and a healing.’’ (Al-hilali M, 1969) (p604) ‘‘And when I am ill, it is He who cures me” (Al-hilali M, 1969) p709. "Never will we be struck except by what Allah has decreed for us; He is our protector" And upon Allah let the believers rely” (Al-hilali M, 1969) (p 904).

While the COVID-19 outbreak keeps spreading, our results will provide important implications for establishing a mental support program to help general population in different regions in Saudi Arabia affected by the COVID-19 epidemic. While the epidemic progresses, it is necessary to prepare healthcare professionals and the general public for medical and mental readiness if there is broad transmission in different regions of Saudi Arabia. Our results have both clinical and policy implications. First, health care administrators should classify high-risk groups as high levels of anxiety for early implementation of a mental health program based on socio-demographic information. Socio-demographic data in this study indicate a higher level of anxiety among married people than unmarried people. This result is in line with previous studies which were found married participants to have higher levels of anxiety than unmarried participants (Li et al., 2020). The logical reason for that is the married people fear for their families and also fear for their children’s academic status. The confusion and potential negative effects on academic advancement may have a negative effect on the students’ mental health. Education During the outbreak, Saudi policymakers need to establish web-portals and web-based applications for classrooms or other teaching activities (Zhang, Ho, & Ho, 2014). Second, government and healthcare agencies should provide reliable information on safety during the epidemic to reduce the effect of rumors (Rubin & Wessely, 2020). Higher contentment with the health information earned was linked to lower levels of anxiety associated with the outbreak. To prevent adverse mental responses, the quality of health information received during the outbreak needs to be based upon evidence. Specific information on medications or vaccinations, transmission channels, and data on the number and location of infected cases (e.g., real-time, online interactive monitoring dashboard) have been linked with lower levels of anxiety. Third, the nature of mental therapies, such as cognitive behavioral therapy (CBT), needs to be reviewed to address the general population’s needs during the outbreak. To prevent the spread of infection, CBT should ideally be administered online or over the phone. Since online CBT does not require the presence of mental health professionals such as psychologists, this will assist the general public in Saudi Arabia, as psychologists are limited in number. Since the general population in Saudi Arabia was homebound by government regulations during the epidemic for at least 12 hours a day, behavioral therapy should concentrate on relaxation activities to alleviate anxiety and scheduling activity such as home-based exercise and entertainment to alleviate anxiety in the home setting.

There are some limitations to the study. Given the limited resources available and the time-sensitivity of the COVID-19 epidemic, the researchers adopted the snowball sampling technique. Sampling technique of snowballing was not based on a random sample collection, and the population studied did not represent the present trend of the general population. Additionally, a prospective analysis on the same group of participants after a period would be appropriate. The researchers were not allowed to obtain contact details and personal data from the respondents because of ethical standards on anonymity and confidentiality. As a result, we could not perform a prospective analysis that would support the need for a focused public health program. Another limitation is that self-reported anxiety of levels may not always be associated with the mental health professionals’ evaluation. This study’s findings may be used as a historical reference point. More significantly, the results specifically advise the creation of mental support programs that can reduce anxiety during COVID-19 epidemic and provide a context for evaluating prevention, control, and treatment efforts during the duration of the COVID-19 outbreak, which is still ongoing at the time of preparation of this study.

## Conclusion

During Saudi Arabia’s COVID-19 outbreak, respondents rated their level of anxiety as mild to moderate. Married respondents reported a higher level of anxiety than unmarried respondents. A lower level of anxiety has been associated with specific, updated and reliable health information and certain preventative measures. The results of this study can be used to develop mental supportive programs during the COVID-19 outbreak to improve mental health and decrease anxiety rates.

## Data Availability

Raw data were generated at [mohannad Alkwiese]. Derived data supporting the findings of this study are available from the corresponding author [initials] on request

